# Strategies for COVID-19 vaccination under a shortage scenario: a geo-stochastic modelling approach

**DOI:** 10.1101/2021.04.19.21255745

**Authors:** N. L. Barreiro, C. I. Ventura, T. Govezensky, M. Núñez, P. G. Bolcatto, R. A. Barrio

## Abstract

In a world being hit by waves of COVID-19, vaccination is a light on the horizon. However, the roll-out of vaccination strategies and their influence on the pandemic are still open problems. In order to compare the effect of various strategies proposed by the World Health Organization and other authorities, a previously developed SEIRS stochastic model of geographical spreading of the virus is extended by adding a compartment for vaccinated people. The parameters of the model were fitted to describe the pandemic evolution in Argentina, Mexico and Spain to analyze the effect of the proposed vaccination strategies. The mobility parameters allow to simulate different social behaviors (e.g. lock-down interventions). Schemes in which vaccines are applied homogeneously in all the country, or limited to the most densely-populated areas, are simulated and compared. The second strategy is found to be more effective. Moreover, under the current global shortage of vaccines, it should be remarked that immunization is enhanced when mobility is reduced. Additionally, repetition of vaccination campaigns should be timed considering the immunity lapse of the vaccinated (and recovered) people. Finally, the model is extended to include the effect of isolation of detected positive cases, shown to be important to reduce infections.

## Introduction

Since the early appearance of SARS-CoV-2 in November 2019 in Wuhan, the world faces a new disease, COVID-19. All governments, without exception, had to quickly devise strategies against this unexpected threat to public health. Meanwhile, the scientific community is advancing in the understanding of all aspects of this disease, which has become a pandemic at a fast rate and with a strength unknown in contemporary history (i.e. more than 127 million cases and 2.7 million deaths worldwide in 16 months,^1^). Some countries are going through the effects of a second or third wave of infections while others are still suffering the first. As the top priority is to diminish the number of susceptible people, in order to minimize the propagation of the virus, several laboratories started to develop vaccines last year. Many of them have reached promising advances and the first generation of vaccines has been already validated by health authorities in many countries.^2,3^

At present, various emergency-approved vaccines have reached an efficacy above 90% in preventing COVID-19 infections, and more importantly, they protect against serious cases requiring hospitalization and reduce lethality.

As a consequence, different vaccination plans and strategies have emerged around the world. More than one year after the isolation of the virus^4^ some people have already been vaccinated (about 320 million people, i.e. *∼*4.1 % of the world population, have received a single dose of vaccine at least, the proportion depending on the country^5^).

Nevertheless, at present vaccine shortage prevails, and international requests are made to the laboratories to increase their vaccine production. More recently, new variants of SARS-CoV-2 with different mutations were detected (e.g. in UK^6^, Brazil, SouthAfrica^7^), which are more transmissible than the previously circulating strain of the virus.

This raises questions about the efficacy of the first set of approved vaccines against the new strains. In any case, this reinforces the need to minimize the propagation of the virus as fast as possible, since contagion favors the appearance of mutations and eventually also of vaccine-resistant strains.

Due to the novelty of the problem, many open questions remain. A critical point to be considered is the immunity period of time provided by the different developed vaccines. It is not clear yet if the immunity lasts a few months, or if it could reach a year. Therefore, even if the problems regarding production, commercialization, distribution logistics, legal aspects,etc. were solved, the schedule or strategies of vaccination are not closed issues.

The complexity of the problem deserves to be treated multidisciplinarily. In this sense compartmental models have been useful to describe and predict different disease scenarios^8^. In particular for countries with large territories and heterogeneous population distribution, such as Mexico and Argentina, geographical spread of the disease must be considered. This family of classical compartmental models can be useful tools only if a degree of local stochasticity is added^9,10^, accounting for the social behavior (mobility, familiar relationships, etc.)^11^. In this work, we have extended previous approaches by including the possibility of a decrease in the number of susceptible people, as a consequence of a schedule of massive vaccination.

Concretely, we use a SEIRS (Susceptible, Exposed, Infected, Recovered, Susceptible) stochastic model of geographical spreading of the disease^10^ and added a compartment to account for vaccinated people, which during the vaccine immunity-period are removed from the susceptible group. We fit the model parameters describing the pandemic evolution in Argentina, Mexico and Spain, respectively, and we analyze the effect of various vaccination strategies with different number of stages and timing regarding their application.

These strategies are based on realistic proposals by the World Health Organization (WHO) and governments, and account for different social behaviors (as lock-down limitations on mobility e.g.), and also if the vaccines are applied either homogeneously or limited to the most densely-populated areas.

Finally, we also included the effect of quarantines on detected positive cases, an effective measure used to reduce infections in many countries, in order to improve the description of the pandemic evolution^11^ and thus assess the long-term vaccination effects more realistically.

Our analyses focus on three different countries (Argentina, Mexico, and Spain), however, the approach could be applied to any country or local jurisdiction that needs to devise a vaccination plan.

In the next section we present the model, then we exhibit and analyze our results, and finally we discuss them, remarking their relevance in order to optimize the effect of vaccination plans in the context of vaccine shortages.

### Model

The present model includes the virus spreading at two levels: (i) Local dynamics: consisting of a compartmental model whose constant parameters are related to the specific disease agent and the host’s immune response, and (ii) Global dynamics of geographical disease spreading: involving mobility parameters related to social habits of the country and affected, at different times, by different non-pharmaceutical interventions by the different governments.

In order to implement the model, a geographical map of the region of interest is divided into square cells with coordinates (*i, j*), covering the whole region. Roads between cities were used to allow short or long distance travelling within the grid. Population size (*N*) is assumed to be constant during the simulated period. If life expectancy is given by *L* and for the mortality we assume an exponential functional form with constant rate *µ* = 1*/L*, then the birth rate should be equal to *µN*.

Population density heterogeneity is considered by using a matrix whose entries are the actual population densities (*ρ*(*i, j*)) inside each cell (at position (*i, j*))

The local dynamics within each cell is calculated using a model with four different compartments (see Fig. 1). These include the susceptible individuals (*S*(*i, j*)) that might become exposed (*E*(*i, j*)) yet not infectious. They become infected (*I*(*i, j*)) after the incubation period *ε* and remain in that state for *σ* days. Afterwards they become recovered (*R*(*i, j*)) individuals, remain immune for *ω* days and close the cycle by becoming susceptible again according to the survival parameter *S*. Vaccines are applied to susceptible people according to a vaccination rate *v*_*r*_. Considering that vaccines arrive in batches, this value is adjusted in each study in order to guarantee the proper administration of all available vaccines in each lot. It is assumed that individuals in the vaccinated (*V* (*i, j*)) compartment have already received as many shots as recommended by pharmaceutics and have developed immunity, thus do not transmit the infection.

**Figure 1.**
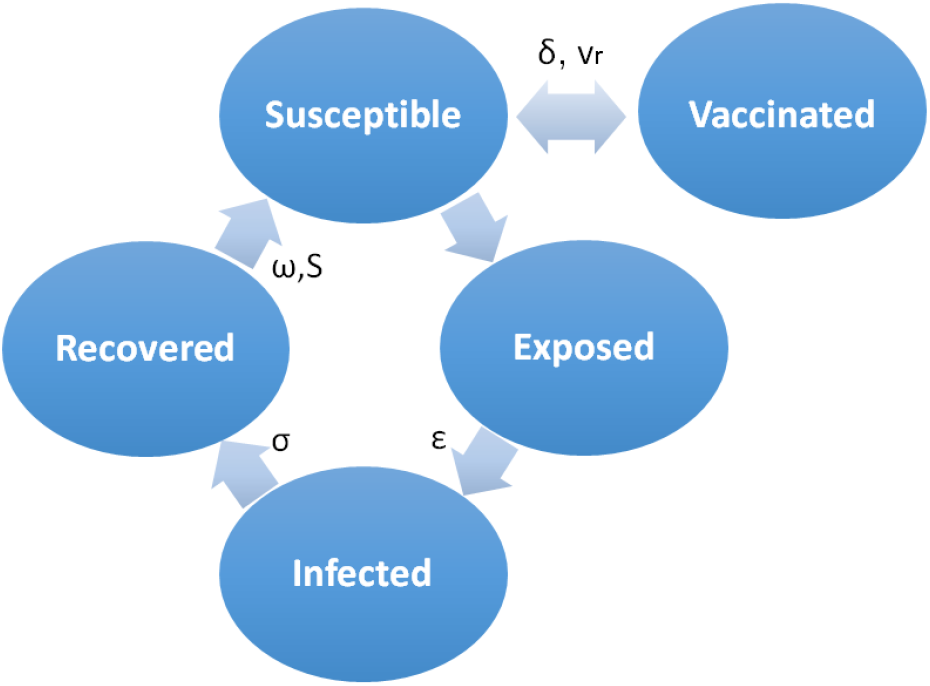
Compartment scheme of a SEIRS-V model. *ε, σ* and *ω* are the latency, infectiousness and immunity periods, *v*_*r*_, *δ* and *S* are the vaccination rate, the vaccine immunity period and the survival parameter, respectively.

However this immunity lasts *δ* days and people become susceptible again after that period. Vaccines are considered 100% effective which is a strong assumption. All time parameters are constant and dimensionless, expressed in a time scale of one day. Based on these assumptions the model is expressed by the discrete mathematical map detailed in eqs. 1-5.

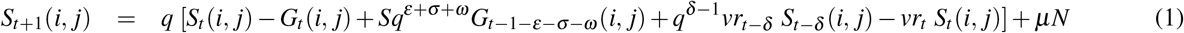

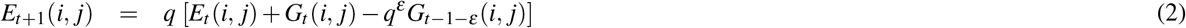

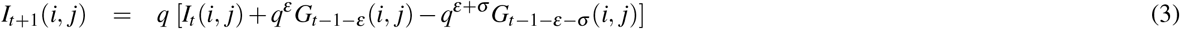

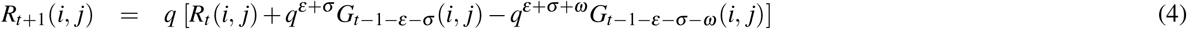

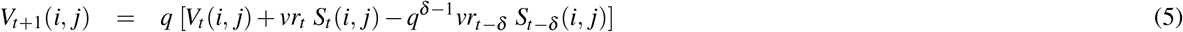

The sum *N*(*i, j*) = *S*(*i, j*) + *E*(*i, j*) + *I*(*i, j*) + *R*(*i, j*) + *V* (*i, j*), is normalized to 1 at t=1. The incidence function 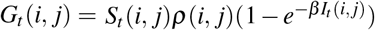 is based on a Poisson probability distribution, assuming a homogeneous mixing within every cell, and taking into account the population density of the specific cell (*i, j*). The transmission parameter *β* is constant and does not depend on population density or mobility but it is a characteristic of the specific pathogen.

It is considered that people sometimes move locally in non-predictable ways. Therefore, randomness is added to cell dynamics. A local mobility parameter per cell (0 *< ν*_*L*_ *<* 1) is compared with a random number from a flat distribution (*r*); if *ν*_*L*_ *≤ r* the epidemic proceeds, otherwise no new infected cases accumulate in that cell due to low mobility during that day.

### Global dynamics

Three mobility mechanisms cause geographical spreading of the virus: by displacements to neighbor cells, by long distance travelling, and by seemingly random trips causing the spread of the disease to remote areas.

Virus spreading to neighbors is often modeled as a diffusion process. An alternative approach is used here by assuming a stochastic process with average cell to cell mobility 0 *< ν*_*n*_ *<* 1 using a Metropolis Monte-Carlo algorithm. Cells considered as possible spreaders are defined with *I*_*t*_(*i, j*) *> η, η* being a parameter related to the infectiousness of the disease. For example, considering (*i, j* + 1) as neighbor cell then if *ν*_*n*_ *≤ r* becomes infected then *S*_*t*_(*i, j* + 1) = 1 *− η* and *I*_*t*_(*i, j* + 1) = *η*.

Long distance travel takes place by air and land thus we define a long distance mobility parameter 0 *< ν*_*a*_ *<* 1. A Metropolis Monte-Carlo algorithm is run again to define if the disease is propagated from cell (*i, j*) to cell (*m, n*). The flows of people between large cities are greater than between small ones. Therefore, in this case *ν*_*a*_*ρ*(*i, j*)*ρ*(*m, n*) is compared to *r*. As before, spreader cells are those where *I*_*t*_(*i, j*) *> η*.

We assume noise in the transmission process because people make non routinary moves in a seemingly random way, allowing for the appearance of epidemic outbreaks in unexpected places. Considering this noise as analogous to the “kinetic energy” of the system, if *e*^*−*1*/KT*^ *> r* and *ρ*(*i, j*) *>*T, (with T being a normalized population density threshold), then the disease is started in the cell (i,j) by making *I*_*t*_(*m, n*) = *η* and *S*_*t*_(*m, n*) = 1 *− η*.

Numerical calculations were performed using maps of population density and road connections for the various countries. (see Supplementary Information: Fig. S1) The size of the grid was of the order of few Km^2^. Since in this model parameters associated with the pathogen and host’s immune response are independent of population density and of people’s mobility, we used *β* = 0.91, *ε* = 1, *σ* = 14 and *ω* = 140, as in R. A. Barrio et al.^10^, and N.Barreiro et al.^11^, for all countries. For simplicity, mobility parameters are considered equal *v*_*a*_ = *v*_*l*_ = *v*_*n*_ = *v*. The parameter *v* changes along time according to non-pharmaceutical interventions adopted by each specific government at specific dates.

### Parameter space exploration

The present shortage of vaccines worldwide highlights the need to propose coherent and lasting strategies to eradicate the pandemic. The model above allows to simulate and analyze different scenarios and administration schemes, in order to determine the parameters that are most relevant to stop the spread of the disease.

With this objective in mind we chose three countries as model systems, namely, Argentina, Spain and Mexico. These countries were chosen because they allow to compare specific aspects of the model. For instance, Argentina and Spain have a similar populations (45 and 47 million, respectively) but they are spatially distributed in a different way. While Argentina has a large territory with few overpopulated areas surrounded by territories with few people, Spain has a more homogeneous distribution of its population. This leads to quite different connections among cities (flow of travellers).

Argentina and Mexico are populated quite heterogeneously but they have very different total populations: 127 and 45 million respectively.

These features are ideal to compare the effect of the model parameters, such as mobility or the increase in the number of vaccines, in different contexts.

In order to analyze their influence in the pandemic evolution, we considered different vaccination strategies by varying a) The total number of vaccines administered in different stages according to their availability; b) the timing between stages; c) the vaccine immunity time, *δ*, which is yet uncertain; d) the social behavior (reflected on people,s mobility) once vaccination begins, and e) the distribution of vaccines, by assuming that the vaccines are applied homogeneously across the country, or vaccinating only in the most densely populated areas. We assume that the efficiency of the vaccines is 100%. If the vaccines were less effective, in this model it would be equivalent to considering that the number of administered vaccines was lower.

At present, in most of the world the number of vaccines is lower than needed. Therefore, it is not possible to ensure their availability, according to the plans made by each government, and many delays are expected. Furthermore, there are multiple variables (political, economical and social) that affect social behavior making it difficult to predict future mobility in each country. Because of this, we emphasize that we do not intend to make specific predictions (although a concrete application of the model would allow it) but to compare different vaccination scenarios presently under consideration by the corresponding decision makers.

### Immunity period and distribution strategies

Due to constraints on the availability of vaccines in the great majority of countries, different vaccine distribution schemes have been proposed (see a review by O.J.Wouters et al,^12^).

To avoid a repetition of the uneven access to vaccines in the H1N1 influenza pandemic of 2009, in April 2020 the World Health Organization announced the creation of the Covid-19 Vaccine Global Access Facility (COVAX), joined already by two-thirds of the world,s countries. To protect the majority of the world,s population against the virus spread and its global impact, COVAX was launched with the initial aim of having 2 billion global vaccine doses available by end of 2021, and a distribution in stages. To ensure equitable access to the world,s largest and most diverse portfolio of safe and effective vaccines, each participant country can request vaccines to cover between 10 and 50% of its population, though will not receive above 20% before all other interested countries received that percentage.

Studies support such a global equitable scheme (see e.g in Chinazzi et al.^13^), predicting a 50% reduction in the number of global deaths could be obtained if the Covid vaccines were distributed proportionally to each country,s population, instead of reserving two-thirds of the vaccines for higher-income countries.

Furthermore, considering the expected initial very limited vaccine supply, two initial stages were suggested by the WHO to reach that 20% coverage of each country,s population: a first stage covering 3%, followed by 17%. Here, we decided to simulate three vaccination stages: those first two WHO-suggested ones, adding a third stage covering 40% of the population, in order to reach a total of 60% of the population vaccinated (closer to the expected values for COVID herd immunity).

Since the vaccine immunity period *δ* is yet unknown, three values of the parameter were assessed, varying the time between stages accordingly. Fig.2 shows the results for the case of Argentina. In Strategy 1: the first stage starts at day 300 after the appearance of the first COVID case, second stage at day 330 and third stage at day 390. In Strategy 2: the first stage starts at day 300, second stage at day 390 and third stage at day 480. Two different geographic distributions of the vaccines are shown: homogeneous, and limited to the most densely populated cities. In all scenarios mobility remains constant from day 300 onward.

Figure 2 evidences that *δ* affects the timing of a new COVID outbreak and also the value of the minimum in the number of daily cases (incidence).

**Figure 2.**
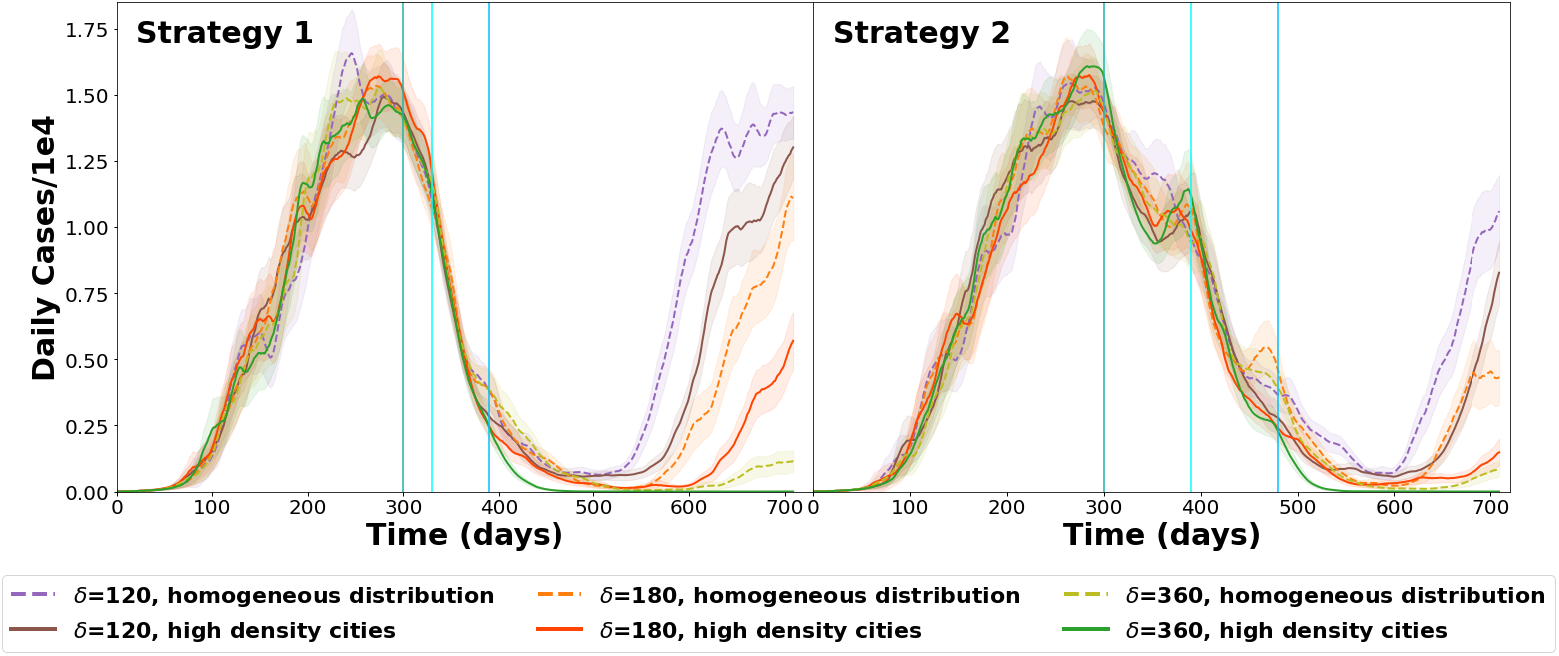
Argentina: Dynamics of daily cases, general results for different values of vaccine-induced immunity period (*δ*). Results for different periods between stages and vaccine geographic distributions. Vertical lines indicate the start of each vaccination stage. The shaded areas correspond to the standard error of each curve.

Incidence decline depends on the timing between stages, in particular, shorter times between stages are more efficient in lowering the total number of cases (see Table S3 in the Supplementary Information). If vaccines are not available for longer periods, so that the time between vaccination stages is longer, the incidence decline is slower and small rebounds may be observed. Although the new outbreak, up to day 720 seems higher for Strategy 2 (see supplementary information Table S3).

Our results show that in case of vaccine shortage, applying doses to the most densely populated areas is more effective, since the total number of cases decreases faster. Furthermore, the minimum number of daily cases reached is lower than when vaccinating homogeneously in the country. The effect is clearly observed for Strategy 1 (further information in Additional Results section of the Supplementary Information).

### Vaccination coverage

To further investigate the effect of the total number of vaccines administered, calculations with Strategy 1 (described above) were performed. An homogeneous distribution of vaccines was simulated while the number of vaccines applied was changed at the third stage from 10% to 50% to obtain a total final coverage of 30% to 70%. Two values of *δ* were used. The results are summarized in Fig. 3.

**Figure 3.**
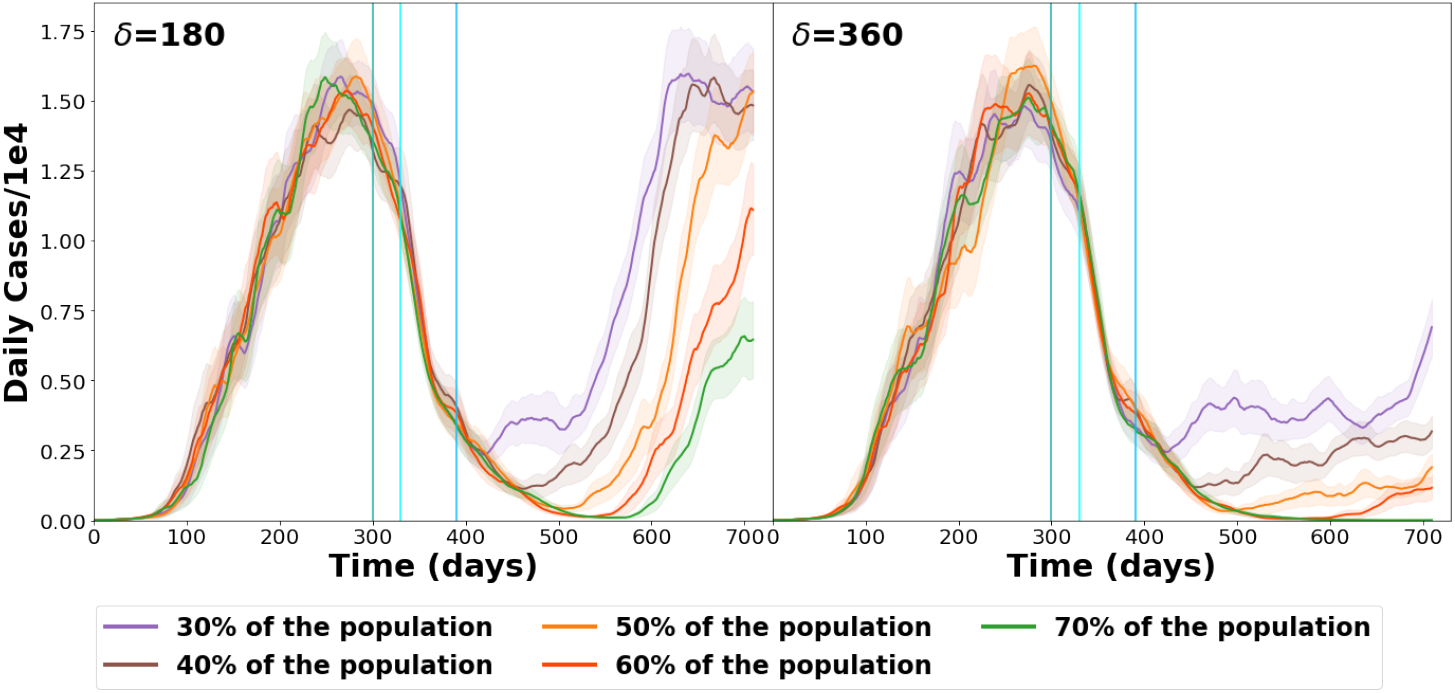
Dynamics of daily cases, general results for different vaccination coverage, for two *δ* values. Vertical lines indicate the start of each vaccination stage. The shaded areas correspond to the standard error of each curve.

For the case of *δ* = 180 days, even with 70% of the population vaccinated, a second wave is observed. However, if only 30% of the population is vaccinated, the incidence lowers, but a steady low incidence period is not observed. In all cases, a second wave starts as the number of immune people decreases, even when 70% of the population is inoculated. When 30%, 40% or 50% of the population is vaccinated, the number of daily cases on day 720 reaches the same levels observed at the maximum of the first wave.

If *δ* = 360 days, the lowest incidence level reached depends on the percentage of the population vaccinated. However, in most cases a slow but steady incidence increase is observed, leading to the appearance of a second wave. However, in all cases a slow but continuous increase in incidence is observed, giving rise to the appearance of a new wave. Only when vaccinating 70% of the population the number of daily cases remains very low (approximately 10 cases per day) all thru to the end of the simulation.

### Impact of social behavior

Social behavior is reflected in the mobility parameters used in the model to fit the reported data for a country. Fig. 4 shows the evolution of the pandemic according to different aspects of vaccination and social behavior in Argentina and Spain. All simulations assume that the vaccines are distributed in 5 stages, as detailed in the upper panel of Table 1. Three curves are compared in each graph: 1)homogeneous vaccine distribution, 2) vaccines applied only in cities with population density above a threshold (see maps with cities included in Fig. S1 in the Supplementary information), and 3) a baseline without vaccination.

**Table 1.** Distribution of vaccines to 62% of the population in Argentina and Spain (upper table). Distribution of vaccines to the whole population of Mexico (lower table)

| # of stage | Period | Argentina vaccines (millions) | Spain vaccines (millions) |
| --- | --- | --- | --- |
| 1st | January | 0.3 | 0.31 |
| 2nd | February | 5 | 5.2 |
| 3rd | March - April | 5 | 5.2 |
| 4th | May - July | 9 | 9.37 |
| 5th | August onward | 8.7 | 9.06 |

| # of stage | Period | Mexico vaccines (millions) |
| --- | --- | --- |
| 1st | February | 0.125 |
| 2nd | March | 17.1 |
| 3rd | April | 13.7 |
| 4th | May | 16.5 |
| 5th | June onward | rest of the population |

**Figure 4.**
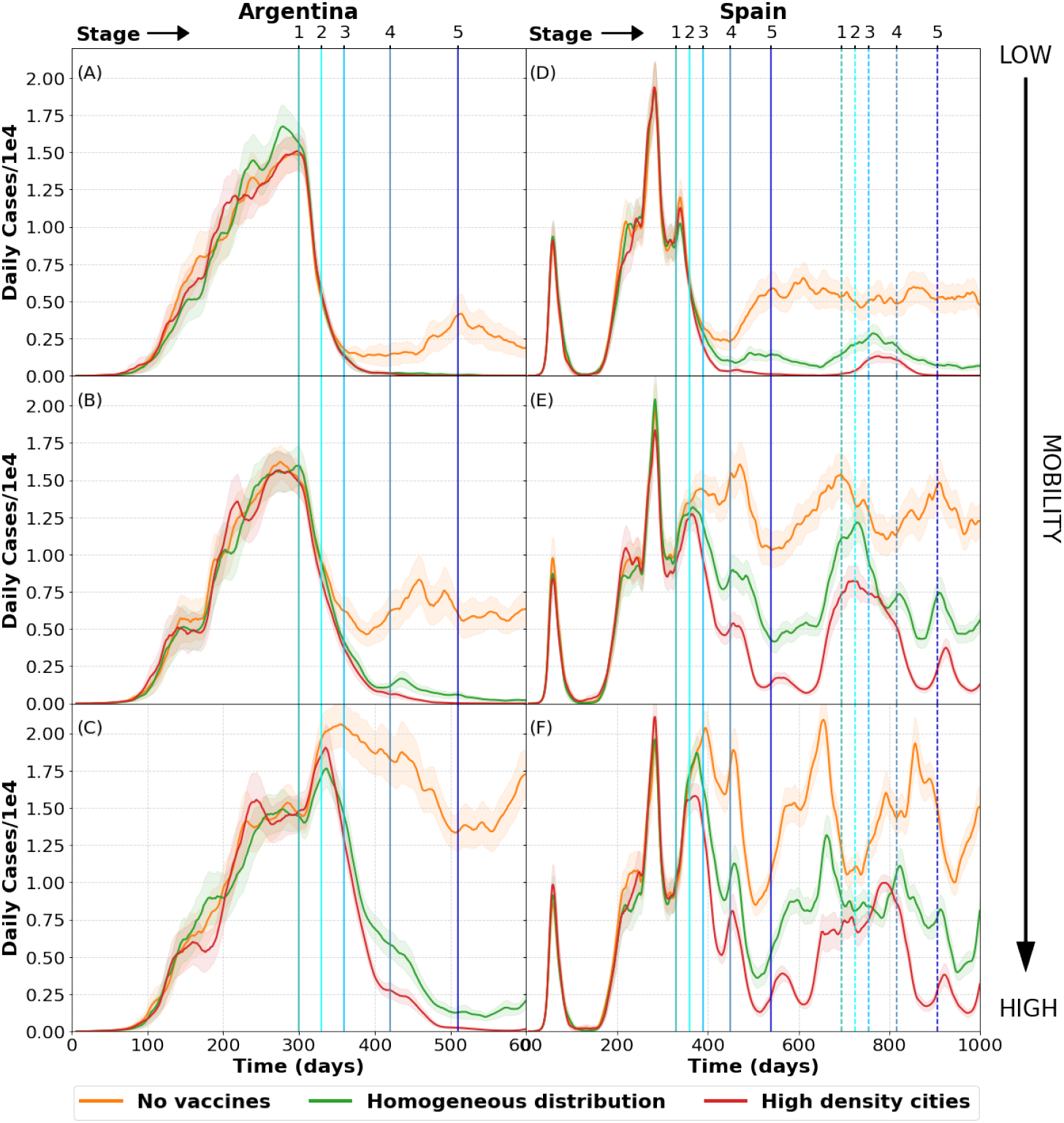
Pandemic evolution in Argentina (left) and Spain (right) according to social behavior. Upper figures ((A) and (D)) show the expected number of infected persons in a low mobility scenario; while figures ((B) and (E)) correspond to a medium mobility scenario, and lower figures ((C) and (F)) to a high mobility scenario. In each graph we consider 3 possible vaccine distributions: None (upper curve), homogeneous, and vaccines applied only in densely populated areas (lowest curve).

Figure 4(A) shows the pandemic evolution in Argentina for the case of low mobility. It simulates a strict lock-down starting in January 2021. It can be observed, as expected, that the amount of daily cases is reduced after a few months. In this case there is no difference between vaccine distribution methods as both of them reduce the amount of daily cases to a minimum. However, due to economic and social stress, it should not be expected that this low mobility could be sustained for a long time. Thus other two situations were modeled, namely with medium and high mobility.

Figure 4(B) shows the evolution of daily cases when social distancing measures are somewhat relaxed. In this case there is a noticeable difference between providing vaccines or not. Moreover, while a homogeneous distribution of vaccines seems to drastically diminish the amount of daily cases, the strategy that reduces the infections to zero is based on prioritizing highly populated areas.

Figure 4(C) shows the model simulations with high mobility, representing the case for no social distancing measures at all. In this situation there is a time lag between the application of the vaccines and their effect, observed on the amount of daily cases, as they keep increasing for a while.

The vaccination of people in highly populated areas is clearly a better strategy to reduce the amount of daily cases This is because most of the infectious hot spots appear in densely populated areas. From there people carry the disease to smaller cities and towns. Moreover, when the main infection sources are turned off, the disease tends to disappear in the whole country.

From these considerations it can be concluded that good results could be achieved for Argentina by applying a strategy combining social distancing and vaccination, even if only 62% of the population were immunized, This percentage corresponds to 28 million people, which is the maximum number of vaccines that might be purchased in Argentina during the whole 2021 year, according to the approved national budget (see Fig. S3 in Supplementary Information).

A similar analysis was performed for the case of Spain modeling analogous vaccination strategies under different representative mobilities (see upper panel in Table 1). Although in this case the strategy is modified in order to vaccinate the same percentage of people at each stage, as it is the case for Argentina (covering 62% of the total population).

Figures 4 (D), (E) and (F) show the daily cases in Spain for low, medium and high mobility respectively. There are three curves in each graph, representing the different vaccination schemes: no vaccination, homogeneous vaccination, and vaccination prioritizing the densely populated cities. All three figures confirm that delivering vaccines in the most densely populated areas is the most efficient method to reduce the number of daily cases.

However, the strategy of distributing doses to only 62% of the population is not as effective in Spain as it is in Argentina for reducing the daily cases. Mobility has a more important role in the virus spread in Spain, as the graphs show. This could be understood in terms of geographic extension and intercity connections: While Argentina has a larger territory and many small isolated cities, Spain is more densely populated and very much connected, in average. Therefore, further outbreaks spread faster throughout the Spanish territory making it harder to stop the pandemic. This points to the conclusion that herd immunity is not only related to the amount of vaccines given but also to other characteristics of the country, for instance, geographic extension, connections, social behavior, etc.

Notice that in Figures 4 (D), (E) and (F) a second vaccination scheme was included for year 2022 (a span of 720 days after the first infected case was reported). Since the immunity of the vaccine is assumed to be of 180 days, a rebound of cases is expected in early 2022 due to the loss of vaccine-immunity. Because of this, a periodical immunization scheme should be considered essential in order to reduce the impact of the pandemic. More examples stressing this point will be shown in the next section.

### Periodical vaccination scheme

As the pandemic seems to be significantly reduced after an adequate relation between responsible social behavior and vaccination, a new question emerges. How long will it last? In order to answer this question we propose different scenarios in Argentina and Mexico that are shown in Fig. 5. The vaccination strategy for Argentina is the same as in table 1. For Mexico we took into account the strategy announced by the government in November 2020. In this case vaccines will be given to the whole population in progressive steps (like e.g. discussed by WHO^14^, or L.Matrajt et al.^15^).

**Figure 5.**
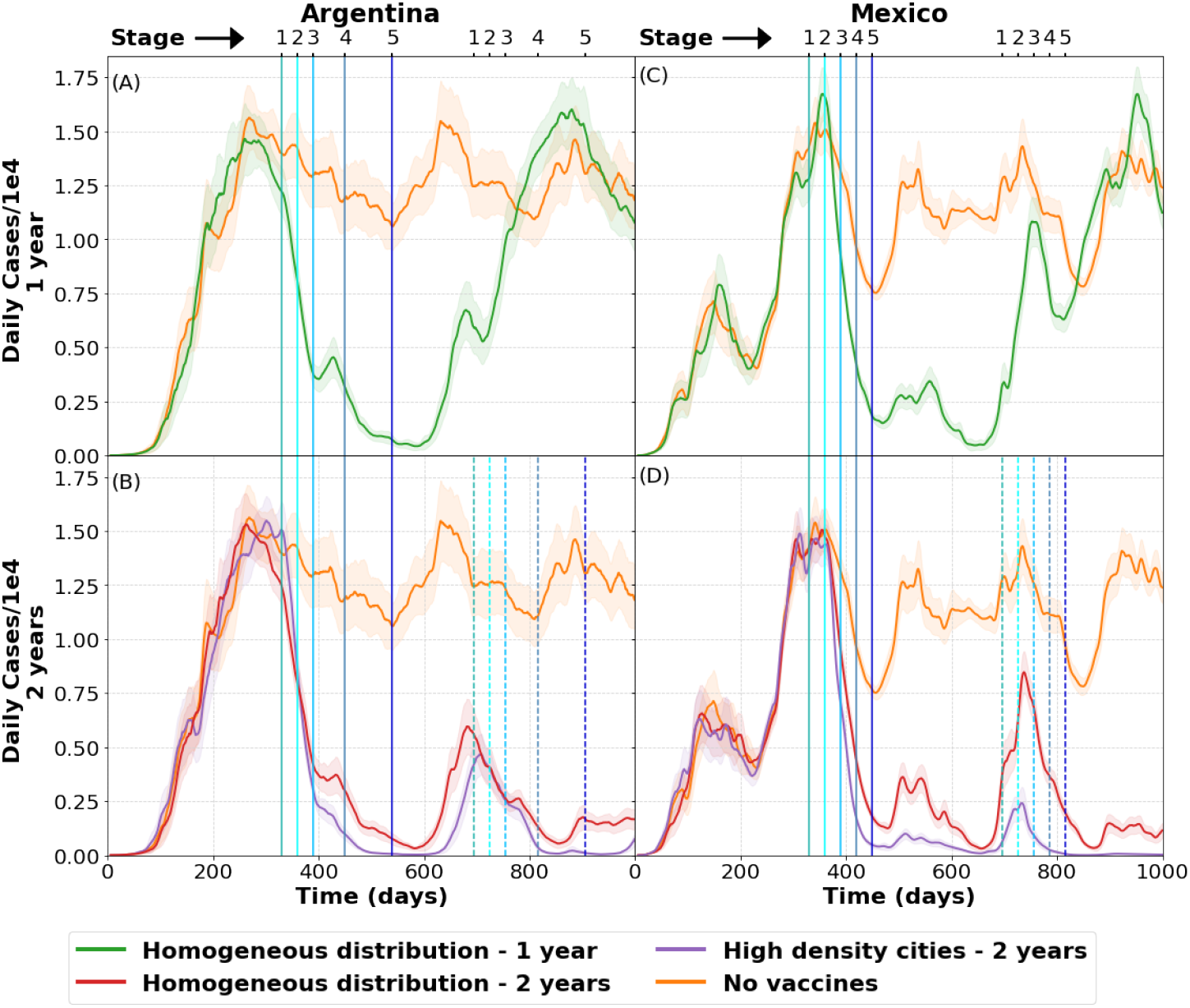
Pandemic evolution in Argentina (A and B) and Mexico (C and D) according to distribution of vaccines. Figures (A) and (C) show the daily cases if the vaccines are applied only during 2021 and the vaccination schemes are not repeated during 2022. In order to compare the results with the cases expected for the regarded mobility, a curve without vaccination (upper curve in each graph) is also included. Figures (B) and (D) show the effect of adding a new vaccination scheme during year 2022. Here we also confirm that vaccination in the most densely populated areas is an additional strategy to reduce the case count. The shaded areas correspond to the standard error of each curve.

First, Mexico will vaccinate health workers, then, people aged more than 60 years. Next, people between 50 and 59, after them people between 40 and 49, and finally, the rest of the population. Taking into account the proportion of the population considered in each age group, we estimated the vaccination strategy shown in the lower panel of Table 1.

Figures 5(A) and (C) show 2 curves: a baseline without vaccines, and homogeneous distribution of vaccines for a scenario where vaccines are given only during 2021. Since in our simulations we assumed that the vaccines provide an immunity period of 180 days, it is expected that the virus will start to spread again and yield a new peak in 2022, reaching the levels of the no-vaccination curve. This behavior is expected to be similar for any vaccine distribution method.

Figures 5(B) and (D) show 3 curves: a baseline without vaccines and two types of distributions, homogeneous or vaccination only in more densely populated areas. In this scenario vaccines are distributed as in Table 1 during 2021 and 2022 giving place to a significant reduction in the second peak for both countries. It should be noticed that the appearance of this second peak is strongly related to the vaccine immunity period, *δ*. A longer period would imply a smaller peak, as far as the same strategy is repeated the next year. But if no vaccines are given the second year, the increment in the daily cases will occur anyway at some point in the future.

As a conclusion and independently of the actual value of *δ*, it is clear that vaccines should be administered to the population periodically to avoid the long term virus spreading. Again, the application of vaccines in highly populated areas gives better results than an homogeneous distribution of doses.

### SEIQRS-V model

Previous studies^16–19^ have shown that most countries are capable of detecting only a fraction of the infected cases. The reason being that most people are asymptomatic or present mild symptoms^20,21^ and do not always seek for medical attention^22^.

Many articles have emphasized the need to improve infected tracking and contact tracing, as a way to reduce the incidence of the pandemic by isolating those detected as positive^23–27^. Taking this into account, track and trace policies, along with social distancing and vaccination, should be considered the main pillars to eradicate the pandemic.

With this in mind, we added a new compartment to our model in order to consider infected people who were isolated. The model with the quarantined (Q) compartment is described in detail in Barreiro *et al*.^11^.

In this work we assume that only a fraction *p* of people who is actually ill is tested and that they get isolated after *α* days of being infected. In this way, this *p* fraction of people stop infecting others reducing the pandemic evolution rate (For more information see Supplementary Information: SEIQRS-V Model section).

It is important to keep in mind that, in this model, government official data for infected cases coincide with the quarantined persons, *i*.*e*. we assume that all the people who tested positive, are in fact isolated and counted as ill by authorities.

In the present work we applied this model to Argentina, Spain and Mexico. As in the SEIRS-V model discussed above, we used the same values for *β, ε, σ* and *ω* for all the countries. *p, α* and *S* where estimated from official data and previous studies. The mobility parameter was fitted in function of time according to changes in governments policies and social behavior (see Supplementary Information: table S2).

Vaccination strategies used in this case can be found in Tables 1 and 2. In all the cases shown, the same vaccination strategy is repeated in 2021 and 2022. The immunity period of the vaccine, *δ*, was conservatively fixed at 180 days.

**Table 2.** Distribution of vaccines in Spain (70% of the population vaccinated until the summer, according to the objectives of the Spanish government^28^). We delayed the strategy one month, to consider the time needed for actual immunization. The shaded areas correspond to the standard error of each curve.

| # of stage | Period | Amount of vaccines (millions) |
| --- | --- | --- |
| 1st | February | 2.3 |
| 2nd | March | 4.5 |
| 3rd | April | 6.6 |
| 4th | May | 6.6 |
| 5th | June | 6.6 |
| 6th | July | 6.6 |

### SEIQRS-V model comparison

In this section we compare the results of different vaccination strategies for the three countries considered.

Figure 6 shows the SEIQRS-V model predictions. Day 1 in the figures coincides with the date of the first reported COVID-19 case in each country. In the Johns Hopkins University database^1^ these dates are March 3rd, February 1st and February 27th for Argentina, Spain and Mexico, respectively.

**Figure 6.**
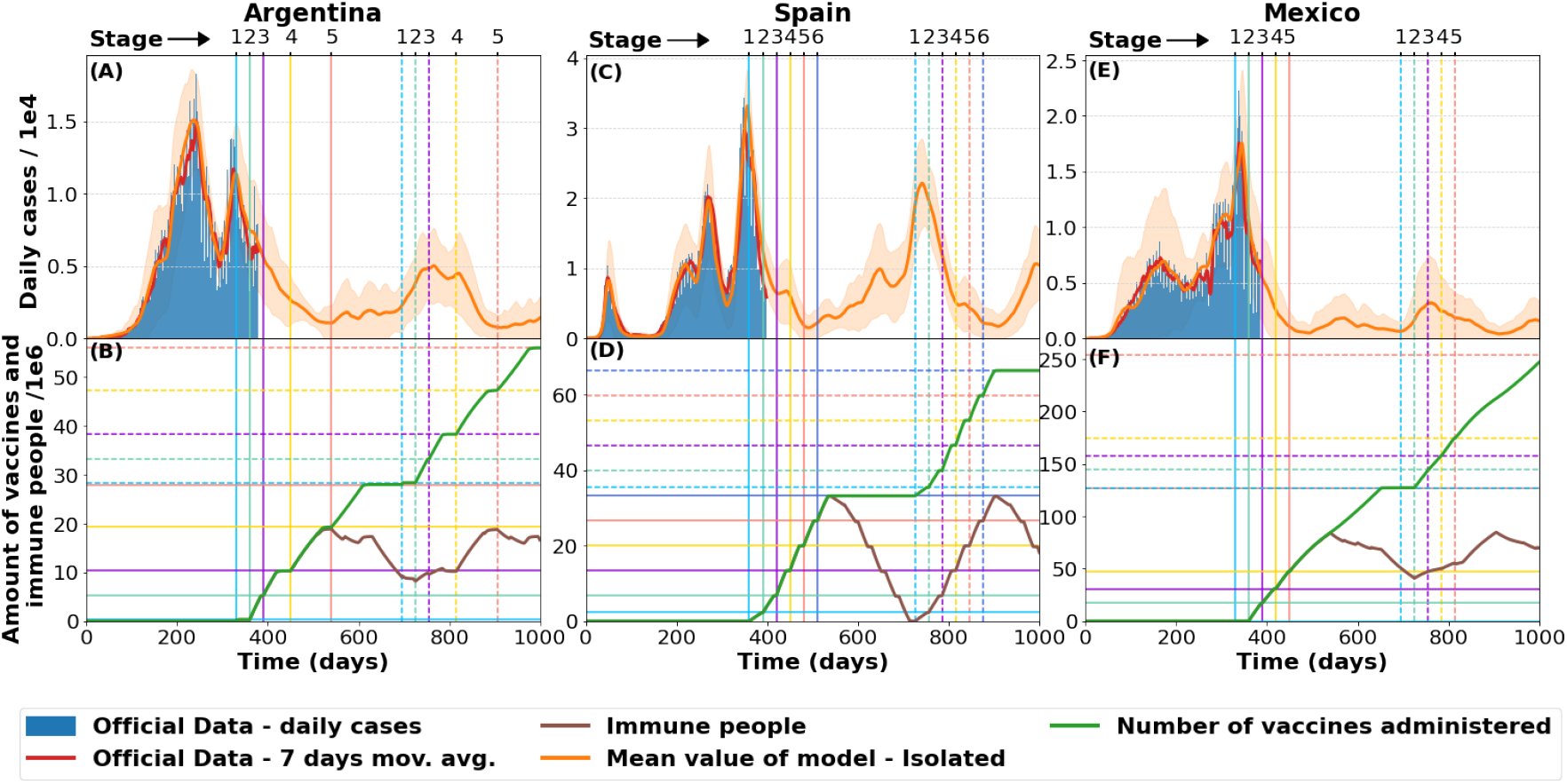
SEIQRS-V model for pandemic evolution in Argentina,Spain and Mexico according to distribution of vaccines.Data updated as of March 7, 2021 (A) Model prediction for Argentina, overlapped with 7 day average of actual daily cases. (B) Immunized people and vaccination strategy for Argentina according to distribution of vaccines on Table 1. (C) Model prediction for Spain overlapped with 7 day average of actual daily cases. (D) Immunized people and vaccination strategy in Spain according to distribution of vaccines (see Table 2).(E) Model prediction for Mexico overlapped with 7 day average of actual daily cases. (F) Immunized people and vaccination strategy in Mexico according to distribution of vaccines (see Table 1). Solid and dashed lines respectively correspond to first and second vaccination year. Horizontal lines in the lower Fig. indicate the available amount of vaccines for each vertical stage (of equal color and aspect).The shaded areas correspond to the standard error of each curve.

For the case of Argentina, mobility was fitted up to day 315 keeping a high value from this moment onward. We also assumed *p* = 0.1 and *α* = 5. The vaccination plan is the same as in Table 1 but delayed for 30 days.

Figure 6 (A) show the number daily cases. One should notice that even with isolation and vaccination, the model predicts that daily cases will never go below 1000 for high mobility. This implies that it is important to implement a better test and trace mechanism or to vaccinate a higher percentage of the population.

It should also be remarked that, in this case, we are considering a relatively small value for vaccine immunity (180 days) and this generates the appearance of a second wave around March 2022. As was shown above for the SEIRS-V model, a longer vaccine immunity period delays the appearance of new pandemic waves.

Figure 6 (B) shows the vaccination strategy as a function of time, and the amount of immune vaccinated people. For a *δ* value of 180 days the amount of immune people starts descending in November 2021, thus causing a growth of daily cases. When a new batch of vaccines is administered in 2022, the amount of immune people increases reducing the impact of the new wave during this year. Under the established immunity conditions, shifting the second vaccination periods to November 2021 could prevent the emergence of this new wave.

Figures 6 (C) and (D) shows the SEIQRS-V model applied to Spain. In this case we used *p* = 0.2, *α* = 5 and a high mobility from the day 343 onward. The vaccination strategy is shown in Table 2 and applied in 2021 and 2022. Figure 6 (C) shows the model prediction together with the reported daily cases. If 70% of the population is vaccinated during the first half of 2021 a strong reduction of daily cases is expected for the summer. Nevertheless, assuming a short immunity period for the vaccine (180 days), the daily cases start growing again generating a new peak around February 2022. This effect could also be understood by looking at Fig. 6 (D). This plot shows vaccinated and immunized people as a function of time. Since vaccines are administered within a short time frame, most people lose immunity by the end of the year. This creates a time window with a low number of immune people that allows new outbreaks and the appearance of a new epidemic wave.

Finally, Figures 6 (E) and (F) show the SEIQRS-V model applied to Mexico. In this case we used *p* = 0.08, *α* = 7 and a high mobility parameter value from day 325 onward. The vaccination strategy is shown in Table 1 and applied in 2021 and 2022.

This strategy establishes that vaccines are administered at an approximately constant rate throughout the year. Therefore a baseline of immunized people is created, which causes the reduction of the susceptible group by almost 50 million people. This kind of administration method also reduces the incidence of new pandemic waves making a difference with respect to the cases of Argentina and Spain.

### Final Remarks

Different vaccination strategies for the Covid-19 were analyzed using a novel epidemiological model with stochastic dynamics of infections in a geographical region. This approach has proven to exhibit a reliable long term predictive power^10,11^.

The parameters defining the disease, as transmission rate, latency and infectiousness periods, can be adjusted with real data taken during a short time after the first outbreak. Furthermore, the parameters regulating the spread of the pandemic in a country have a clear and direct relationship with the measures taken by the authorities and followed by citizens.

This strategy has allowed us to predict the dynamical behavior of the pandemic in three very different countries, like Argentina, Mexico and Spain. There, we include the suggested vaccination strategies and analyze the results under different scenarios which take into account the difficulties caused by the current global vaccine shortage.

The mobility parameters of our model allow to simulate different social behaviors (e.g. lock-down interventions, travel restrictions, social distancing), and analyze the effect of different vaccine distribution strategies. One could also analyze the predictions using a strategy based on a homogeneous vaccination of the population, or when prioritizing the most densely populated areas.

In a context of doses shortage, our model predicts that the most effective strategy in reducing infections is the vaccination of the most-densely populated areas.

Since the epidemic reappears periodically, we found that vaccination campaigns should be repeated. In order to optimize the effect of these, one should take into account the immunity period of the vaccines to prevent premature future outbreaks. Keeping the number of contagions low is of great importance, as a large number of infections favors the appearance of mutations and, eventually, of new vaccine-resistant strains.

Regarding the so called herd immunity, understood as the successful suppression of the epidemic reached when enough people are immune to the virus, previous works estimate its threshold for SARS-CoV-2 in the range from 10% to 70% or more. The lower values might not hold true because of the assumptions made about people interactions in social networks^29^. However, for the case of COVID-19, we learnt from our model that it depends on the number of already infected people and the interplay of disease and vaccine related-factors. Among the virus factors we considered the influence of the immunity time after recovery and the probability of reinfections. While for the vaccine-related factors, we explored the effects of the time period of immunity conferred by the vaccine, the percentage of population vaccinated, and the pace of vaccine delivery.

The probability of infection is affected by logistical, social and demographic factors and it is key to the adopted vaccination strategy. As was mentioned before, there are huge differences between a scheme based on a demographic homogeneous vaccine delivery and one that prioritizes the most densely-populated areas. We found that the vaccination impact is larger in countries with less connected cities, combined with a low mobility.

From modelling concrete vaccination strategies in countries with very different demographic features, it can be clearly concluded that in the case of a limited number of vaccines, herd immunity is not achieved using vaccination strategies alone. A combination of mobility control, detection and isolation of infected people (and probably social distancing measures) is needed to drastically reduce infections. Such measures have negative socio-economic impacts at the local (and global) level. However, these could be minimized by optimizing the allocation of vaccines.

Some guidelines can be learnt from our model as it was shown and discussed in this work. However, the worldwide vaccination allocation should be approached carefully.^30^ There are 130 nations with 2.5 billion people that have not received a single vaccine yet^31^, where the COVID-19 virus with uncontrolled transmission, has a perfect playground to evolve into dangerous variants, even immune to the developed vaccines.

An outbreak there, has the potential to become an outbreak anywhere. In this respect, our model could be extended from one particular country to a continent or even to the whole world in order to compare different vaccine global distribution strategies.

Despite the real threat, the current global vaccine allocation is driven by national interest and consequently it is highly asymmetric. High-income countries, representing 16% of the world’s population, purchased more than half of all COVID-19 vaccine doses^31^ available to date, aside the COVAX agreement. This will extend the time frame of the pandemic and the economical burden worldwide^13,32,33^. Unless a worldwide vaccine allocation strategy based on health and epidemiological needs is implemented, humanity will not be able to achieve global herd immunity soon enough and will suffer the consequences of COVID-19 for much longer.

## Data Availability

OK

## Acknowledgements

RAB, MN and NLB acknowledge support from The National Autonomous University of Mexico (UNAM) and Alianza UCMX of the University of California (UC), through the project included in the Special Call for Binational Collaborative Projects addressing COVID-19. RAB was financially supported by Conacyt through project 283279. MN was partially supported by CONICET. We appreciate useful discussions with Prof. Yang Quan Chen and collaborators.

## Author contributions statement

N.L.B., C.I.V., T.G. and R.A.B. conceived the idea. N.L.B. proposed and wrote the model in python, created the maps and did the fitting for each country. N.L.B, C.I.V., T.G. and M.N. conducted the simulations. All authors discussed the results, wrote and reviewed the manuscript.

