## Supplementary material for "Strategies for COVID-19 vaccination under a shortage scenario: a geo-stochastic modelling approach": Supplemmentary information

### Additional information on methods

#### Information to run simulations of the model

As was mentioned before, in order to run the model code, it is necessary to have a population map and the main road connections inside each country. Each map is divided in a grid of square cells in order to consider micro and macrodynamics of the disease. The information from Argentina was obtained from the IGN (Instituto Geográfico Nacional). The population information from Spain and Mexico was obtained from worldpop[1] and the connections were obtained from Google Maps. With the retrieved information we generated the maps from fig S1. In the case of Argentina and Mexico we divided the map in a grid of 7 x 7 $km^2$ cells. The smaller area and higher density of Spain allowed to separate its map in a smaller grid with 5 x5 $km^2$ cells The three maps on the left, S1(A), S1(C) and S1(E) show the density maps overlapped with the connections used to run the model. The maps on the right S1(B), S1(D), S1(F) represent the areas with higher population used in some vaccination strategies described in the main text.

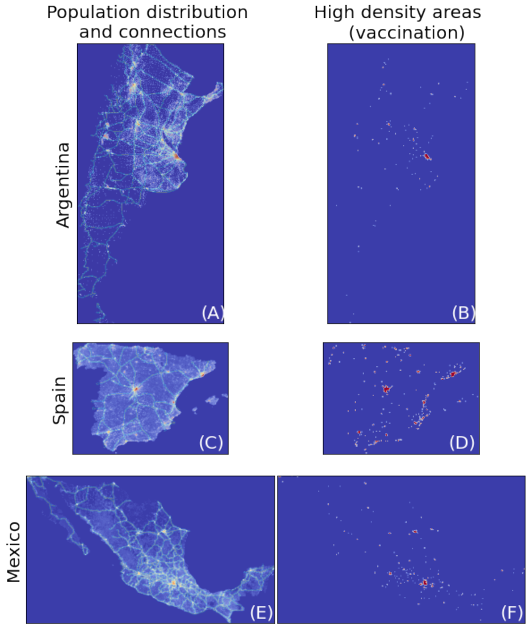


**Figure S1.** Maps used to run the model. Figures (A), (C) and (E) are the population distribution and connection maps used in Argentina, Spain and Mexico respectively to run the code. Figures (B),(D) and (F) exhibit the densely-populated areas where vaccines are applied.

The $\eta$ parameter, which is related with the infectiousness of the disease, was fixed to different values according to population density and grid used in each country. We used $10^{-5}$ , $10^{-4}$ and $5\,10^{-6}$ for Argentina, Spain and Mexico respectively. In all the cases it corresponds to starting the disease in a city with around 10 infected people. The survival parameters $S$ used for each country can be found in table S1. Final noise and mobility parameters are detailed in the next section.

### SEIQRS-V Model

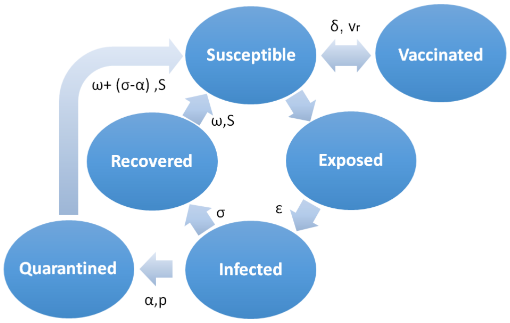


**Figure S2.** Compartment scheme of a SEIQRS-V model. $\varepsilon$, $\sigma$ and $\omega$ are the latency, infectiousness and immunity periods, $vr$, $\delta$ and $S$ are the vaccination rate, the vaccine immunity period and the survival parameter, respectively. $p$ is the fraction of infected isolated people and $\alpha$ the time period from infection to isolation.

In this case the SEIRS-V compartment model was replaced by a SEIQRS-V model where $Q$ stands for quarantined people. Stochastic geographical spread features are the same as those described before. Two new variables were added to consider the fraction of people discovered and isolated ($p$) and the time period between infection and isolation ($\alpha$). A schematic graph of this model can be seen in figure S2. Equations S1-S6 show the addition of this new compartment to the SEIRS-V model. In this case we consider that people tested positive for Covid-19 are not vaccinated until they lose their natural immunity $\omega$.

$$S_{t+1} = (1-\mu)\,(S_t - G_t + S(1-\mu)^{\varepsilon+\sigma+\omega}\,G_{t-1-\varepsilon-\sigma-\omega} + (1-\mu)^{\delta-1}\,vr_{t-\delta}\,S_{t-\delta} - vr_t\,S_t) + \mu N \tag{S1}$$

$$E_{t+1} = (1-\mu)\,(E_t + G_t - (1-\mu)^{\varepsilon}\,G(t-1-\varepsilon)) \tag{S2}$$

$$I_{t+1} = (1-\mu)\,(I_t + (1-\mu)^{\varepsilon}\,G_{t-1-\varepsilon} - (1-p)\,(1-\mu)^{\varepsilon+\sigma}\,G_{t-1-\varepsilon-\sigma} - p\,(1-\mu)^{\varepsilon+\alpha}\,G_{t-1-\varepsilon-\alpha}) \tag{S3}$$

$$Q_{t+1} = (1-\mu)\,(Q_t + p\,(1-\mu)^{\varepsilon+\alpha}\,G_{t-1-\varepsilon-\alpha} - p\,(1-\mu)^{\varepsilon+\sigma+\omega}\,G_{t-1-\varepsilon-\sigma-\omega}) \tag{S4}$$

$$R_{t+1} = (1-\mu)\,(R_t + (1-p)\,(1-\mu)^{\varepsilon+\sigma}\,G_{t-1-\varepsilon-\sigma} - (1-p)\,(1-\mu)^{\varepsilon+\sigma+\omega}\,G_{t-1-\varepsilon-\sigma-\omega}) \tag{S5}$$

$$V_{t+1} = (1-\mu)\,(V_t + vr_t\,S_t - (1-\mu)^{\delta-1}\,vr_{t-\delta}\,S_{t-\delta}) \tag{S6}$$

#### Model fitting

The parameter $p$ was estimated from bibliography and local surveys from each country. For instance, a study by Figar et al.[2] suggests that only 10% of the infected are discovered in Argentina. A zero prevalence study in Spain by Pollan et al.[3] indicates that the $p$ parameter should be between 0.1 and 0.3. In our case we were able to fit the data using $p = 0.2$. Finally, an ongoing survey from ENSANUT[4] suggest a high presence of COVID-19 antibodies in the Mexican population, implying a small $p$ value.

**Table S1.** Parameters used in each simulation

| Country | $p$ | $\alpha$ | $S$ |
|---|---|---|---|
| Argentina | 0.1 | 5 days | 0.9973 |
| Spain | 0.2 | 5 days | 0.9973 |
| Mexico | 0.08 | 7 days | 0.9919 |

The value $\alpha$ was estimated from official data in each country. Assuming that a person can be contagious at least 48 hours before presenting symptoms, we added 2 days to the time between appearance of symptoms and actual testing. Nevertheless, for small values of $p$, variations in the time between infection and isolation have low impact in the results[5]. Table S1 shows the values of $\alpha$ and $p$ used in each case. The table also shows the survival parameter used in each case. We estimated the dates related with changes in mobility and overall social behaviour from two main sources. The first one was the government intervention dates related with restriction of population mobility and implementation of social distancing policies. This information was acquired from the Oxford stringency index[6]. This index measures the strictness of the government policies taken to face the pandemic on time. The other source was the COVID-19 Community Mobility Reports from Google LLC[7] Table S2 shows intervention times and mobility parameters fitted to each country.

**Table S2.** Mobility and noise parameters fitted for each country. Around day 300 all the countries show an increase of mobility because of the holidays

| | Argentina | | | Spain | | | Mexico | |
|---|---|---|---|---|---|---|---|---|
| **Intervention day** | **Mobility** | **Noise** | **Intervention day** | **Mobility** | **Noise** | **Intervention day** | **Mobility** | **Noise** |
| 0 | 0.33 | 0.1 | 0 | 0.46 | 0.1 | 0 | 0.73 | 0.1 |
| 23 | 0.135 | 0.1 | 32 | 0.23 | 0.1 | 16 | 0.183 | 0.1 |
| 79 | 0.188 | 0.1 | 39 | 0.06 | 0.1 | 60 | 0.161 | 0.1 |
| 102 | 0.223 | 0.1 | 101 | 0.08 | 0.1 | 92 | 0.319 | 0.3 |
| 166 | 0.356 | 0.1 | 130 | 0.12 | 0.1 | 174 | 0.412 | 0.3 |
| 235 | 0.28 | 0.1 | 143 | 0.2 | 0.1 | 225 | 0.42 | 0.5 |
| 285 | 0.51 | 0.1 | 152 | 0.3 | 0.1 | 255 | 0.58 | 0.5 |
| 315 | 0.356 | 0.1 | 169 | 0.24 | 0.1 | 305 | 1 | 0.9 |
| – | – | – | 180 | 0.2 | 0.1 | 325 | 0.42 | 0.3 |
| – | – | – | 209 | 0.18 | 0.1 | – | – | – |
| – | – | – | 234 | 0.42 | 0.1 | – | – | – |
| – | – | – | 258 | 0.18 | 0.8 | – | – | – |
| – | – | – | 301 | 0.37 | 0.3 | – | – | – |
| – | – | – | 313 | 0.96 | 0.8 | – | – | – |
| – | – | – | 343 | 0.54 | 0.8 | – | – | – |

#### Code

The model was developed in python using two libraries: NumPy[8] and Pandas[9]. Plots were created with Matplotlib[10]. All the curves were obtained by averaging 100 runs of the model. The model code and the archives used to run simulations will be made available upon request.

### Additional Results

#### Vaccination shortage and Argentina National Budget for 2021

Though the vaccination in Argentina is proceeding at slower pace than planned (around 2.98 million people received at least one dose by 29 March,[11]), due to failure to obtain the vaccines from laboratories at the originally expected rate, taking into account the national budget for the government approved by the Congress in Argentina for year 2021, we can ascertain the effect of the vaccination program. Concretely, the 2021 national budget has envisaged an expenditure for Covid vaccine purchases amounting to a minimum of 11 million and a maximum of 28 million vaccines.

In Figure S3 we show the effect of the minimal and maximal vaccination schemes foreseen by the 2021 national budget, if they were applied in 3 main stages. In the case of the minimal budget: the two first stages give place to an important reduction of the number of infections (to about a third of the main peak of infections), but still an important number of cases would persist during 2021 (a minimum of about 4000 daily cases exists, if vaccines were applied homogeneously in the whole country, or of 2500 daily cases if vaccination was applied in the most densely-populated cities), with an increase of cases during the last trimester.

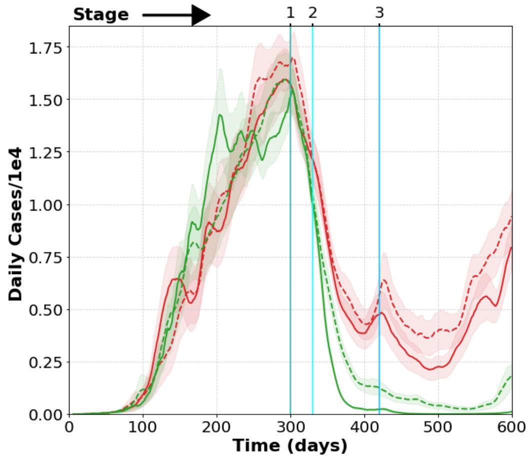


**Figure S3.** Argentina National budget for 2021: minimal-11 million ( red) and maximal-28 million ( green) total vaccine purchases. Effect of 3-stage vaccination on number of daily infections: vaccination stages starting at 300, 330, and 420 days (vertical lines) from first infected case (March,3th 2020). Assumed: immunity time of recovered patients is 140 days, and vaccine immunity time is 180 days. Solid lines: cases if vaccination prioritizes densely-populated areas. Dashed lines: cases if vaccines are homogeneously distributed throughout the country.

Figure S3 also shows the effect of the maximal vaccination scheme. In this case, a very important reduction of the number of infections is obtained with the two first vaccination stages: now to about an eighth of the main peak of infections, reaching a much smaller number of cases afterwards during 2021 (a minimum of about 500 daily cases exists, if vaccines were applied homogeneously in the whole country, and a negligible number of daily cases would be reached if vaccination was applied on the most densely-populated cities), with a small increase of cases during the last trimester. Our results prove that, especially in a context of vaccine shortage, it is clearly advantageous to focus the vaccination on the cities with higher population density in order to more effectively reduce the propagation of the virus. Also, that the maximal vaccination scheme approved in the national budget (reaching about 62 percent of the population) would be required in order to reach a strong reduction of virus propagation in Argentina in 2021.

### Additional information on immunity period and distribution strategies

The results presented in Fig 2 in the main text are clear, however some details are better illustrated in Table S3. The upper table shows the accumulated cases at the end of the simulation, equivalent to 720 days of pandemic. Accumulated cases are less when vaccinating high population-density areas in comparison to vaccinating homogeneously throughout the country. For $\delta = 120$, the previous result is observed regardless of the timing between stages (Strategy 1 vs Strategy 2); but for $\delta = 180$ or $\delta = 360$ the total number of cases is lower for Strategy 1 than for Strategy 2. The number of daily cases at the minimum steady incidence period (between days 485 and 540 for Strategy 1, and between days 575 and 630 for Strategy 2), was averaged (lower table). Minimum averages decrease as $\delta$ increases. Notice that for $\delta = 360$, using Strategy 1 and vaccinating in high population-density areas, incidences of about 5 cases per day are achieved, making it feasible not only to isolate infected people, but to track their contacts so they can be isolated too.

**Table S3.** Total number of accumulated cases (upper table). Minimum number of daily cases (lower table)

| | Strategy 1 | | Strategy 2 | |
|---|---|---|---|---|
| $\delta$ | Homogeneous | High Density | Homogeneous | High Density |
| 120 | 4,739,832 | 4,084,429 | 4,535,998 | 4,062,800 |
| 180 | 3,758,801 | 3,212,385 | 4,090,868 | 3,695,505 |
| 360 | 3,070,143 | 2,813,648 | 3,779,243 | 3,605,921 |

| | Strategy 1 | | Strategy 2 | |
|---|---|---|---|---|
| $\delta$ | Homogeneous | High Density | Homogeneous | High Density |
| 120 | 722 | 634 | 979 | 659 |
| 180 | 248 | 203 | 288 | 319 |
| 360 | 270 | 5 | 146 | 137 |
